# Socio-demographic characteristics and their relation to medical service consumption among elderly in Israel during the COVID-19 lockdown in 2020 compared to the corresponding period in 2019

**DOI:** 10.1101/2022.01.29.22269933

**Authors:** Ohad Shaked, Liat Korn, Yair Shapira, Avi Zigdon

**Author notes:** **Corresponded author** Prof. Liat Korn, Ph.D., Kiryat HaMada St 4, Ariel, Email address, Address: Ariel University, Science Park, P.O.B. 3, Ariel, 40700, Israel. Kiryat HaMada St 4, Ariel, Email address, Address: Ariel University, Science Park, P.O.B. 3, Ariel, 40700, Israel. (W), Address: Ariel University, Science Park, P.O.B. 3, Ariel, 40700, Israel.

## Abstract

**Purpose:** The COVID-19 pandemic has led to the isolation of the population in Israel, including the elderly, some of whom are chronically ill. The present study examines the consumption of medical services among adults over the age of 65 in Israel at the time of the first COVID-19 lockdown relative to the corresponding period the year before, according to various socio-demographic variables: gender, age, marital status, ethnicity, degree of religiosity and socioeconomic status.

**Methods:** A retrospective longitudinal observational quantitative research based on the “Natali Healthcare Solutions Israel” database of subscribers. Company subscribers over the age of 65 (N=103,955) were included in the sample (64.5% women) in two time periods, before the COVID-19 outbreak-P1, in 2019, and during the first COVID-19 lockdown-P2 in 2020. Medical needs included referrals to a medical or emergency services, ordering an ambulance, physician home visits and service refusal.

**Results:** The average number of referrals to services was lower during the COVID-19 lockdown period (M=0.3658, SD=0.781) compared to the corresponding period the previous year (M=0.5402, SD=0.935). At the time of the COVID-19 lockdown, the average number of referrals to medical services was lower, but the average number of ambulance orders, physician visits and service refusals was higher compared to the same period in the previous year. During both time periods, women (P1-M=0.5631, SD=0.951; P2-M=0.3846, SD=0.800) required significantly more (*p<*.*000*) services than men (P1-M=0.5114, SD=0.910; P2-M=0.3417, SD=0.753). In both time periods, subscribers who were older, widowed, living in non-Jewish/mixed localities or in average or below average socioeconomic status localities required more services relative to younger, married people living in Jewish localities, or in above-average socioeconomic localities (*p<*.*000*).

**Summary and Conclusions:** In a large sample of elderly in Israel, findings indicate a decrease in referrals to medical care during the first COVID-19 lockdown period, yet an increase in ambulance orders, physician visits and service refusals. There were no significant differences between the periods according to different socio-demographic characteristics. The period of the first COVID-19 lockdown was characterized by a higher incidence of medical service refusals compared to routine times. The incidence of requiring medical service at the time of the first COVID-19 lockdown was 2.5 times higher among those who required services during the previous year, 1.7 times higher among unmarried seniors, and 1.4 times higher among the older age group of 85 and over in comparison to the younger seniors.

## Introduction

The COVID-19 pandemic has caught the world population with insufficient readiness, especially with respect to the elderly who are particularly vulnerable to serious health consequences. Travel restrictions, including the prohibition of leaving homes, as well as the fear of contracting the disease have made it difficult for the elderly. Elderly and people who are chronically ill were forced to isolate in order to prevent exposure to disease effects, with the government’s stated goal being that this was done to protect them and other at-risk groups [1]-[4]. The elderly were instructed not to come into work but to work remotely as much as possible, to use delivery services or family members for food and medication purchases and to avoid any unnecessary meetings. It was shown that this age group strictly adhered to the guidelines compared to the general population [1], [4], [5].

Technology has been the heart of adaptation in general and among the elderly population in particular. Telehealth services became the center of the medical field response, with elderly and high-risk patients staying home and being treated remotely [6]. In Israel, some of the response to the elderly during the COVID-19 pandemic was given through technological means that assisted them in coping with the pandemic. These services ranged from medical services to logistics services as well as personal telephone support. Various socio-demographic aspects that characterize the elderly population have been found to be related in their readiness to deal with a global catastrophic event such as the COVID-19 pandemic. The present study examines the effect of the COVID-19 pandemic on service consumption among the elderly in Israel during the COVID-19 lockdown in 2020 compared to the corresponding period in 2019 by socio-demographic characteristics.

### Socio-demographic variables as risk factors in service consumption among the elderly

Dealing with the activities of daily living of the elderly is greatly affected by their health status [7]. In addition, multiple morbidity, disabilities, and lack of functional independence have been shown to be risk factors for the adaptive daily conduct of the elderly [8], [9]. Another study found that loneliness and age have been found to be risk factors for low adherence to guidelines, in a manner in which the older or lonelier a person is, the lower their compliance is with guidelines. That is, the behavioral routine in old age shows a tendency for less compliance with medical guidelines [10]. Many seniors experience situations of loneliness, high morbidity, disability, lack of independence, multiple medication use and income struggles. These may lead to mental burnout that delay recovery from conditions they are experiencing, as well as a decrease in compliance with medical recommendations, a matter indicating disobedient behavior that can lead to various complications [10]-[12]. On the other hand, as people age, the higher the mortality rate is and as a result, the higher the fear of mortality, especially among elderly with various background diseases [13]. It had also been shown that the demand for emergency care is higher among adults than among young people and higher among Jews (3.1 times) than among Arabs (2.1 times) [14].

With respect to gender in old age, various studies have found that women tend to consume more medical services, adhere more with medical guidelines, attend more medical examinations, and obtain emergency services in higher frequency in comparison to men [10], [15]-[17].

Social support is identified as a strategy that can improve health outcomes [18]. Family members and social support within the family contribute greatly to the management of diseases among the elderly [19]. A person’s family status is found to be related to receiving support in times of crisis, and are a crucial factor in disease management [19], [20]. A married person, or one in a relationship, is assisted by the partner who wakes up with them in the morning, is aware of their changing condition, reminds them to take their medications and the likes. Being without the support of a partner has a negative impact on the management of chronic illness and medical condition. An article by Liao and colleagues found that mutual support between elderly spouses improves lifestyle and reduces complications among chronic patients, especially in the initial prevention phase of disease deterioration [20]. Older people were found to be more likely to require emergency services among married couples than among unmarried partners [21]. Older widowed women tend to ignore their deteriorating health, if they do not suffer from a serious medical condition [22]. There is a relationship between people adhering to values, beliefs and customs based on religion and healthy behavior in old age. It was found that the degree of religiosity influences a person’s decisions, their behavior, and their degree of compliance with medical guidelines. Often, and especially in old age, religiosity is a powerful source of spiritual strength, comfort, and hope, and has an impact on decision-making and medical compliance [23].

A study conducted in the United States following the national emergency subsequent to COVID-19, found that the elderly fear obtaining emergency room care, and a decrease of 72% was observed in the number of visits to emergency medicine clinics. Despite this data, an increase in visits to emergency medicine clinics was observed with respect to symptoms of infectious diseases or mental health problems [24]. Additional studies conducted in the United States have found similar findings which reinforce the fear of getting to the emergency room for medical care. Even patients with acute myocardial infarction - heart attack, arrived less in the emergency room, in a manner which delayed treatment of conditions that may result in mortality without appropriate treatment [25]-[27].

Health disparities between Jewish and Arab patients may stem, among other things, from cultural differences and accessibility to health centers [14]. This may be compared to the highest prevalence of COVID-19 in the USA being observed mainly among African American and Latino ethnic minorities, as well as among immigrants, as these groups are characterized by low socioeconomic status, inaccessibility to medical services, exacerbation of morbidity and complications of the disease, and are at increased risk for exposure to COVID-19 within their workplaces and in their more crowded living situations [28].

In Israel, various organizations provide a wide range of services to the elderly, including medical services such as a doctor’s home visit, ambulance services, and telephone counseling as well as various logistical services such as medication purchasing and hot meal deliveries. “Natali Healthcare Solutions in Israel” is one of the more prominent Israeli companies for providing these services, in which the subscriber contacts the center and speaks to a medic who personally tailors the service required to them. In life-threatening situations, an ambulance will be dispatched to provide immediate medical assistance to the patient, whereas in situations where there is no imminent risk, the call center may supply other services such as telephone counseling or a doctor’s home visit. In terms of logistic services, a wide range of options is available, for instance: laundry services, medication home delivery, and hot meals service. The current study is based on data obtained from “Natali Healthcare Solutions in Israel” during the COVID-19 era and in the preceding year.

The previous pandemic which led to closures in different continents around the world, occurred about 100 years ago during the Spanish flu outbreak [29]. The relationship between socio-demographic characteristics of the elderly population in the context of consumption of medical services has not been sufficiently investigated and there are not enough relevant publications examining this topic since the Spanish flu outbreak. The importance of examining these characteristics among Israel’s elderly population at the time of the first lockdown and its implications on other lockdowns during the pandemic is significant for the purpose of understanding the needs of this population with respect to health promotion, especially when this part of the population is continuously growing in percentage throughout the years. The purpose of the present study is to examine the variability in demand of medical services among elderly in Israel during the first COVID-19 lockdown in comparison to the corresponding period in the previous year, by distribution of various socio-demographic variables, as well as how this demand for medical services is characterized according to different socio-demographic variables: gender, age, marital status, ethnicity, degree of religiosity and socioeconomic status. We hypothesize that service consumption would be lower during the COVID-19 pandemic compared to the corresponding period in the previous year, and that men, widowers, adults in the younger age group as well as non-Jews will consume fewer services in comparison to women, married, older individuals and Jews.

## Methods

This study is a longitudinal observational retrospective study that compares the consumption of medical services by the elderly in two time periods. The first period – prior to the start of the pandemic in February-April 2019 (P1). The second period - the corresponding months in the following year, February-April 2020 (P2), in which the citizens of Israel were subjected to the COVID-19 lockdown restrictions imposed by the government, in accordance with Ministry of Health directives and guidelines.

The study is based on a database from “Natali Healthcare Solutions in Israel”, which is a private company for the provision of medical services that has been operating in Israel for over 30 years. The company provides a wide range of medical and non-medical services to elderly subscribers and is part of a range of organizations operating in the field medical service provision to Israeli residents. The company’s services include telemedicine services supported by call center representatives who have been trained as emergency call medics, as well as medical consultation calls with various physicians, emergency medical services such ambulance services and physicians’ home visit services. As part of the service, the company provides subscribers with medical devices such as: distress buttons, ECG devices, various sensors, and passive monitoring devices [30].

The study was approved by the Ariel University institutional ethics committee (Approval No. AU-HEA-AZ-20200624) and has also received the approval of “Natali Healthcare Solutions in Israel” management to conduct the research. Data from the research file was taken retrospectively and anonymously from the company’s database without identifying details of the subjects. The subjects’ records were kept in an encoded file in a manner which ensured the identification of the subjects stayed concealed and that no link could be made between the data and the individuals sampled.

### Population and sample description

The study population is adults over the age of 65 in Israel who are “Natali Healthcare Solutions in Israel” subscribers. Of the full subscriber list for the years 2019-2020, the following customers who did not meet the study criteria were removed: those under the age of 65, those whose services were terminated for various reasons (cancellation, death, service freeze - 2% of the sample), those whose number of calls to the center was over 400 inquiries in both periods (n=17). The sample size meeting the study criteria was 103,955. Table 1 presents the final sample according to the socio-demographic variables examined.

**Table 1:** Sample description by socio-demographic variables.

| Variable | Values | Total Sample |  |
| --- | --- | --- | --- |
|  |  | n | % |
| **Total Sample** |  | **103955** | **100%** |
| **Gender** | Males | 65954 | 63.5 |
|  | Females | 36364 | 35.0 |
|  | Unknown | 1637 | 1.6 |
| **Age Categories** | Young seniors (65-75) | 30252 | 29.1 |
|  | Mid-age seniors (76-85) | 47846 | 46.0 |
|  | Elder seniors (86 and above) | 25855 | 24.9 |
| **Marital status** | Bachelor | 1105 | 1.5 |
|  | Married | 46360 | 61.6 |
|  | Divorced | 4496 | 6.0 |
|  | Widowed | 23252 | 30.9 |
| **Ethnicity** | Jewish | 87210 | 83.9 |
|  | Non-Jewish | 918 | 0.9 |
|  | Mixed | 15825 | 15.2 |
| **Residential religiosity** | Secular residence | 102305 | 98.4 |
|  | Religious residence | 421 | 0.4 |
|  | Orthodox residence | 1229 | 1.2 |
| **Socioeconomic status of residence** | Below average (clusters 1-5) | 39124 | 37.6 |
|  | Average (clusters 6-7) | 38677 | 37.2 |
|  | Above average (clusters 8-10) | 26152 | 25.2 |

Total of 103,955 “Natali Healthcare Solutions in Israel” subscribers participated in the study, representing the entire customer population for the 65+ age group (Mean age 80, SD=7.46, Range 65-119). Table 1 shows the sample included 64.5% women (n=65,954) and 35.5% men (n=36,364). 1637 of the participants did not have gender information in their files. The average age of the subjects was 80 years (SD=7.46) which we divided into three age categories: 29.1% constituted the young age group-65-75, 46.0% constituted the middle age group-76-85, and the oldest age group of those aged 86 and over accounted for 24.9% of the sample, of whom 61.6% were married and 30.9% widowed. The sample had a Jewish 83.9% and secular 98.4% majority. Over one third of the sample 37.2% reside in localities of an average socio-economic status of 1-5, which is below the Israel Central Bureau of Statistics [31] average, 37.2% within the socio-economic status of 6-7, and 25.2% in the above-average localities 8-10.

### Study Variables

#### Dependent variables

Various medical needs constituted the dependent variables in the study. The dependent variables were measured, for each period separately: before COVID-19 (P1), and during lockdown (P2). <u>Demand for services</u> - For the purpose of measuring the demand for services, a general index was composed of four variables for each period separately. The four variables comprising the index are: medical calls to the center, emergency calls to the center, ordering an ambulance and calling for a doctor’s visit (Cronbach’s alpha at P1 = 0.693, and at P2-0.630). <u>Medical calls to the center</u> - The number of times a subscriber contacted the call center for medical reasons (for example, by phone or via distress button). The value scale in P1 was 0-114, and in P2-0-472. <u>Emergency calls to the center</u> - The number of times a subscriber contacted the emergency call center. The value scale in P1 was 0-226, and in P2-0-294. These two variables consisted of a “total referrals” index that included the count of medical referrals and emergency referrals together for each subject. The value scale in P1 was 0-294, and in P2-0-540. <u>Ordering an ambulance</u> - The number of times the subscriber ordered an ambulance during these periods created a scale of values as detailed: P1 0-18, and P2-0-16. <u>Ambulance cancellation</u> - The number of times the subscriber canceled an ambulance ordered during these periods. The value scale at P1 was 0-7, and at P2-0-6. <u>Doctor’s visits</u> - The number of times a subscriber called a doctor during these periods. The value scale at P1 was 0-102, and at P2-0-111. <u>Service Refusal</u> - The number of times during the period, in which an ambulance service or a doctor’s visit were offered by the call center, but the subscriber refused the service. The value scale at P1 was 0-6, and at P2-0-18. The variable of service refusal was not included within the general variables of service consumption since it expresses a cancellation of needs and was therefore not measured. For the construction of the services consumption index, the four variables were first recoded into dichotomous feasibility variables that received the values of 0 - no service was required, and 1 required service, and then the four were summed into one complex index for each period with a 0-4 value scale.

#### Independent variables

The data file contained the socio-demographic variables of the subjects, which were used as independent variables in this study as detailed: <u>Gender</u> – male / female. <u>Age</u> - calculated at the starting point of the study - 2019 by year of birth. The age of two of the subjects was unknown. A new age variable was created by grouping the ages into three categories [32] as detailed: young seniors (65-75), middle-aged adults (76-85), older adults (86 and older). <u>Marital status</u> - single / married / divorced / widowed. 28,742 subjects (27.5% of the sample) were missing data on marital status.

Three additional socio-demographic variables were derived from the respondent’s residential address after cross-referencing the information with Israel CBS data [31] as follows: Ethnicity, degree of religiosity and the socio-economic status of the residence. Ethnicity <u>of the residence</u> – the residential address of the subscriber was examined via CBS database. The values of the variable are: 1. Jewish locality, 2. Non-Jewish locality, and 3. mixed ethnic locality. <u>The degree of religiosity of the residence</u> - by city of residence. The values of the variable are: 1. a secular locality, 2. a religious locality, and 3. ultra-Orthodox locality. <u>Socio-economic status of the residence</u> - according to CBS coding, the localities are classified into clusters from 1-10, with 1 symbolizing a very low socioeconomic status, and 10 a very high socioeconomic status. This scale was divided into three categories: 1. Low socioeconomic status - clusters 1-5, 2. Middle socioeconomic status - clusters 6-7, and 3. High socioeconomic status - clusters 8-10 [31].

### Data Analysis

For data analysis purposes, the file was centralized in a data table in the IBM SPSS Statistics 25 software. To adapt the data to the senior population in Israel, the data file was weighted by gender and age. The data in Table 1 presents descriptive statistics for the sample by the Frequency command, before and after weighting the data. Missing values were omitted from the description of the variables in Table 1, which displays the valid percent value as well as the raw number value of the subjects in each category. Table 2 presents averages and standard deviations using a T test for paired variables comparing between the two periods. The 2-tailed significance of the T value analysis is presented. Table 3 presents the averages of the index comprised of “service demand” by period. A Two-way mixed ANOVA with Bonferroni correction for pairwise comparisons, was used to examine the differences by group, time, and by the interaction between group and time while examining significance by Bonferroni. Similarly, Table 4 presents the same analysis for the dependent variable of service refusal. Values displayed in Bold - are those significantly higher values than the other groups. Table 5 presents logistic regression results for predicting service demand according to the study’s socio-demographic variables. For this purpose, the dependent variable “service demand” was divided dichotomously in P2 by a median. The β values displayed in the table constitute the Relative risk - RR and present the probability of service demand according to the independent variable groups. The confidence Interval – CI Range displays the minimum and maximum values for the range with a 95% of confidence. Values displayed in Bold - are significantly higher values than the other groups.

**Table 2:** **Mean of service consumption for the elderly population According to study periods – pre COVID-19 (P1) and during COVID-19 lockdown (P2)**

|  |  | **P1** | **P2** | **Significance**<br>**(2 tailed)** | **T value** |
| --- | --- | --- | --- | --- | --- |
| **Total service Demands** | M | 1.194 | 0.756 | 0.000 | 32.86 |
|  | SD | 3.71 | 4.32 |  |  |
| **Ambulance orders** | M | 0.082 | 0.087 | 0.011 | -2.55 |
|  | SD | 0.38 | 0.40 |  |  |
| **Ambulance cancellations** | M | 0.006 | 0.009 | 0.000 | -8.10 |
|  | SD | 0.08 | 0.10 |  |  |
| **Physician's visit** | M | 0.106 | 0.122 | 0.000 | -7.25 |
|  | SD | 0.60 | 0.72 |  |  |
| **Refusal of service** | M | 0.013 | 0.022 | 0.000 | -12.22 |
|  | SD | 0.14 | 0.20 |  |  |

M=Mean, SD= Standard deviation, N=103955

**Table 3:** **Mean of medical service consumption among the elderly population in the pre-COVID-19 period (P1), and during COVID-19 lockdown (P2) by socio-demographic variables**

| Variables / Values |  | P1 |  |  | P2 |  |  | Time | Inter. | N |
| --- | --- | --- | --- | --- | --- | --- | --- | --- | --- | --- |
|  |  | M | SD | Sig. | M | SD | Sig. | sig. | sig. |  |
| **Gender** | Males | **.5631<sup>a</sup>** | .951 | 0.000 | **.3846<sup>a</sup>** | .800 | 0.000 | 0.000 | NS | 65954 |
|  | Females | .5114<sup>b</sup> | .910 |  | .3417<sup>b</sup> | .753 |  |  |  | 36364 |
| **Age Categories** | Young seniors | .3848<sup>c</sup> | .796 | 0.000 | .2389<sup>c</sup> | .640 | 0.000 | 0.000 | <0.001 | 30252 |
|  | Mid-age seniors | .5500<sup>b</sup> | .933 |  | .3734<sup>b</sup> | .776 |  |  |  | 47846 |
|  | Elder seniors | **.7040<sup>a</sup>** | 1.052 |  | **.5001<sup>a</sup>** | .906 |  |  |  | 25855 |
| **Marital status** | Bachelor | .5946<sup>b</sup> | .931 | 0.000 | .4027<sup>b</sup> | .796 | 0.000 | <0.001 | <0.001 | 1105 |
|  | Married | .4571<sup>c</sup> | .884 |  | .2978<sup>c</sup> | .711 |  |  |  | 46360 |
|  | Divorced | .6210<sup>b</sup> | .953 |  | .4446<sup>b</sup> | .826 |  |  |  | 4496 |
|  | Widowed | **.7451<sup>a</sup>** | 1.013 |  | **.5406<sup>a</sup>** | .906 |  |  |  | 23252 |
| **Ethnicity** | Jewish | .5355<sup>b</sup> | .932 | 0.000 | .3630<sup>b</sup> | .779 | 0.000 | <0.001 | 0.052 | 87210 |
|  | Non-Jewish | **.6111<sup>a</sup>** | .994 |  | **.3725<sup>a</sup>** | .741 |  |  |  | 918 |
|  | Mixed | **.5620<sup>a</sup>** | .945 |  | **.3808<sup>a</sup>** | .795 |  |  |  | 15825 |
| **Residential religiosity** | Secular residence | .5398<sup>a</sup> | .934 | 0.097 | .3659<sup>a</sup> | .781 | 0.109 | <0.001 | NS | 102092 |
|  | Religious residence | .4991<sup>a</sup> | .852 |  | .3026<sup>a</sup> | .718 |  |  |  | 575 |
|  | Orthodox residence | .5893<sup>a</sup> | 1.00 |  | .3835<sup>a</sup> | .809 |  |  |  | 1288 |
| **Residential SES** | Below average | **.5547<sup>a</sup>** | .945 | 0.000 | **.3729<sup>a</sup>** | .784 | 0.000 | 0.000 | NS | 39124 |
|  | Average | **.5575<sup>a</sup>** | .941 |  | **.3867<sup>a</sup>** | .798 |  |  |  | 38677 |
|  | Above average | .4929<sup>b</sup> | .909 |  | .3241<sup>b</sup> | .748 |  |  |  | 26152 |
| **Total services consumption** |  | .5402 | .935 | 0.000 | .3658 | .781 | 0.000 |  | - | 103955 |

P1=pre-COVID-19 period; P2=during COVID-19 lockdown; M=Mean; SD=Standard deviation; Sig=Significant; Inter. Sig.=Interaction significant; N=Number of subjects; SES Socioeconomic status Significance testing for differences between groups $a \neq b \neq c \neq d$ where $a$ represents the higher value, $p < 0.05$ .

**Table 4:** **Mean service refusal (ambulance and physician’s visit) among the elderly population in the pre-COVID-19 period (P1) and during COVID-19 lockdown (P2) by socio-demographic variables**

| Variables / Values |  | P1 |  |  | P2 |  |  | Time sig. | Inter. sig. | N |
| --- | --- | --- | --- | --- | --- | --- | --- | --- | --- | --- |
|  |  | M | SD | Sig. | M | SD | Sig. |  |  |  |
| **Gender** | Males | .0156<sup>a</sup> | .152 | 0.000 | .0248<sup>a</sup> | .219 | 0.000 | <0.001 | NS | 65954 |
|  | Females | .0102<sup>b</sup> | .118 |  | .0185<sup>b</sup> | .165 |  |  |  | 36364 |
| **Age Categories** | Young seniors | .0097<sup>c</sup> | .125 | 0.000 | .0193<sup>c</sup> | .221 | 0.000 | 0.000 | NS | 30252 |
|  | Mid-age seniors | .0137<sup>b</sup> | .140 |  | .0220<sup>b</sup> | .190 |  |  |  | 47846 |
|  | Elder seniors | .0178<sup>a</sup> | .157 |  | .0265<sup>a</sup> | .195 |  |  |  | 25855 |
| **Marital status** | Bachelor | .0244<sup>a</sup> | .255 | 0.000 | .0281<sup>a</sup> | .208 | 0.000 | 0.000 | NS | 1105 |
|  | Married | .0111<sup>b</sup> | .125 |  | .0187<sup>b</sup> | .189 |  |  |  | 46360 |
|  | Divorced | .0178<sup>a</sup> | .163 |  | .0269<sup>a</sup> | .199 |  |  |  | 4496 |
|  | Widowed | .0186<sup>a</sup> | .158 |  | .0299<sup>a</sup> | .229 |  |  |  | 23252 |
| **Ethnicity** | Jewish | .0136<sup>a</sup> | .141 | 0.867 | .0224<sup>a</sup> | .196 | 0.758 | 0.003 | NS | 87210 |
|  | Non-Jewish | .0120<sup>a</sup> | .127 |  | .0174<sup>a</sup> | .160 |  |  |  | 918 |
|  | Mixed | .0131<sup>a</sup> | .138 |  | .0224<sup>a</sup> | .229 |  |  |  | 15825 |
| **Residential religiosity** | Secular residence | .0135<sup>a</sup> | .140 | 0.212 | .0224<sup>a</sup> | .201 | 0.932 | 0.019 | NS | 102092 |
|  | Religious residence | .0052<sup>a</sup> | .072 |  | .0191<sup>a</sup> | .171 |  |  |  | 575 |
|  | Orthodox residence | .0186<sup>a</sup> | .170 |  | .0233<sup>a</sup> | .174 |  |  |  | 1288 |
| **Residential SES** | Below average | .0136<sup>a</sup> | .138 | 0.295 | .0215<sup>b</sup> | .194 | 0.013 | 0.000 | NS | 39124 |
|  | Average | .0142<sup>a</sup> | .143 |  | .0246<sup>a</sup> | .221 |  |  |  | 38677 |
|  | Above average | .0125<sup>a</sup> | .139 |  | .0202<sup>b</sup> | .178 |  |  |  | 26152 |

P1=pre-COVID-19 period; P2=during COVID-19 lockdown; M=Mean; SD=Standard deviation; Sig=Significant; Inter. Sig.=Interaction significant; N=Number of subjects; SES Socioeconomic status Significance testing for differences between groups $a \neq b \neq c \neq d$ where-a represents the higher value, $p < 0.05$ .

**Table 5:** **Logistic regression results for the prediction of medical service consumption during COVID-19 lockdown by socio-demographic variables and pre-COVID-19 service requirements**

| Variables | Values | $\beta$ | Confidence interval (CI) | |
| --- | --- | --- | --- | --- |
|  |  |  | Low | High |
| **Gender** | 0=Females, 1=Males | **1.093\*\*\*** | 1.049 | 1.138 |
| **Age** | 0=Younger, 1=Elder | **1.397\*\*\*** | 1.340 | 1.457 |
| **Marital status** | 0=Married, 1=Not married | **1.757\*\*\*** | 1.688 | 1.828 |
| **Ethnicity** | 0=Jews, 1=Non-Jews | 1.001 | .984 | 1.018 |
| **Residential religiosity** | 0=Secular, 1=religion | 1.001 | .922 | 1.085 |
| **Residential SES** | 0=Low and below, 1=average | **1.137\*\*\*** | 1.095 | 1.181 |
| **P1 Services consumption** | 0=No consumption, 1=yes | **2.489\*\*\*** | 2.441 | 2.537 |
| **n** |  |  |  | **74878** |
| **Explained variance** | **Nagelkerke R Square** |  |  | **24.0%** |

\*p<.05, \*\*p<.01, \*\*\*p<.001

### Findings

The data file containing 103,955 records of “Natali Healthcare Solutions in Israel” subscribers in two periods was the platform for analyzes that examined the research questions. First, we examined all service requirements including the total referrals to the center, ambulance orders, ambulance cancellations, doctor’s home visits and the refusal of service. For this purpose, a T-test analysis was performed for the paired averages of the dependent variables. Table 2 displays the differences between the dependent variables in the two time periods.

It can be seen from Table 2 that the averages of all types of service requirements differ between the pre-COVID-19 period (P1) and the COVID-19 lockdown period (P2) significantly. The total number of calls to the center was the only variable that averaged higher in P1 (M=1.194, SD=3.71) in comparison to P2 (M=0.756, SD=4.32) significantly (*p<*.*000*). That is, during P2, there was a lower average of calls to the center relative to P1. Ambulance orders (M=0.087, SD=0.40) and ambulance cancellations (M=0.009, SD=0.10) were higher in P2 than in P1 (M=0.082, SD=0.38; M=0.006, SD=0.08). Similarly, the average number or ordering a doctor’s home visit was higher during first COVID-19 lockdown period (M=0.122, SD=0.72) than during the pre-COVID-19 period (M=0.106, SD=0.60). Also, refusal of service (ambulance or doctor’s visit) was higher in P2 (M=0.022, SD= 0.20) than in P1 (M= 0.013, SD= 0.14).

Table 3 presents the averages of the collective index of service requirements, which was consisted of medical calls to the center, emergency calls to the center, ambulance orders and home doctor’s visits pre-COVID-19 and during the first COVID-19 lockdown period and according to various socio-demographic variables. In order to check if there were any differences between needs in P1 and P2 within study groups we preformed mixed repeated measures.

It can be seen from Table 3 that in total, the average demand for services decreased from the pre-COVID-19 period (M=0.5402, SD=0.935) in relation to the first COVID-19 lockdown period (M=0.3658, SD=0.781). In both time periods, women (P1-M=0.5631, SD=0.951; P2-M=0.3846, SD=0.800) required more services than men (P1-M=0.5114, SD=0.910; P2-M=0.3417, SD=0.753) Significantly (*p<*.*000*). In both time periods, the older age group required more services than the younger age groups, widowers required more services in relation to single, divorced or married individuals; subscribers residing in non-Jewish or mixed localities required more services in relation to those living in Jewish localities significantly; and seniors living in localities socioeconomically rated as average or below average required more services in relation to those living in above average rated localities (*p<*.*000*). No significant differences in the request for services were found according to the degree of religiosity of the locality in the two time periods. Significant interaction effects were found between age groups, family status and ethnicity.

Similarly, to what is shown in Table 3, Table 4 shows the average of ambulance or home doctor visits service refusals, pre-COVID-19 and during the first COVID-19 lockdown period and according to various socio-demographic variables.

It can be seen from Table 4 that in both time periods, women (P1-M=0.0156, SD=0.152; P2-M=0.0248, SD=0.219) refused service more than men (P1-M=0.0102, SD=0.118; P2-M=0.0185, SD=0.165) Significantly (*p<*.*000*). In both periods the older age group refused service more than the younger age groups, and unmarried seniors-single, divorced and widowers refused service more than married seniors significantly. No significant differences were found in the refusal of services by sector or degree of religiosity by locality in the two periods. In P2 the average service cancellation was highest among localities ranked in the average socioeconomic status (M=0.0246, SD=0.221) relative to low socioeconomic status localities (M=0.0215, SD=0.194) or above average ones (M=0.0202, SD =0.178).

Table 5 presents results of logistical regression for predicting service requirements during the first COVID-19 lockdown period from socio-demographic variables and pre-COVID-19 service requirements.

The logistic regression analysis shows that the most powerful variables for predicting services requirements during P2, are demand for services in the previous year (RR = 2.489, p <.000, 95% CI-2.441-2.537), marital status (RR = 1.757, p <.000, 95% CI-1,688-1,828), and age (RR = 1,397, p <.000, 95% CI-1,340-1,457). The variables of gender and socioeconomic status came out significant but not strong in the model. The variables of sector and religiosity did not come out significant in the model. These variables explain 24.0% of the variance of the dependent variable – service requirement during the COVID-19 lockdown period of time. These findings indicate that the chance of requiring service at the time of the first COVID-19 lockdown was 2.5 times higher among those who required services in a previous year, 1.7 times higher among unmarried seniors, and 1.4 times higher among the older age group of 85 and over in comparison to the younger seniors.

## Discussion

The COVID-19 pandemic has greatly affected the elderly population [1], and the various restrictions imposed due to it have brought to refrain from leaving the home for various needs and even for the purpose of receiving medical care [1], [3]. This study characterized the consumption of services among the elderly in Israel during the first COVID-19 lockdown in 2020 compared to the corresponding period in 2019, according to various socio-demographic characteristics,based on data from “Natali Healthcare Solutions Israel” database of subscribers.

This study is unique both in examining topics that have not come up in pervious Israeli studies examining the COVID-19 effects, as well as in its reliance on a very large sample of the elderly population in Israel, which requires medical resources and social support in routine times. Consumption of the various services that “Natali Healthcare Solutions Israel” offers, was examined in this study with reference to inquiries made to the call center – via phone or distress button, ambulance orders, doctor’s home visits as well as refusals to receive service. These variables were analyzed in both time periods while examining their distribution by gender, age, religious sector, degree of religiosity and socio-economic status.

An investigation of the findings according to the study periods shows that during the period of the first COVID-19 lockdown in 2020, the average number of service requirements by elders to “Natali Healthcare Solutions in Israel” various social and medical services was significantly lower compared to the corresponding period in 2019, in line with the study hypothesis. In times of crisis, consumption of medical services is expected to decrease, as found in a study conducted in the USA in which chronic patients turned less to medical service centers [24]. It was found that people avoided going to emergency rooms and at times, even when experiencing life-threatening situations, did not come in for a prompt diagnosis or treatment [24]-[27].

Loneliness among the elderly is associated with less effective disease management, and deterioration is expected to be slower among non-single elderly people due to disease management and their social environment [19]-[21]. It is possible that the lower average of service requests during the first COVID-19 lockdown, as found in the present study, is due to the need for seniors to temporarily move in with their families, thus requiring less external support services. The COVID-19 period and its subsequent restrictions, have raised anxiety levels and fears of the elderly about contracting the disease due to the higher likelihood of dying from it [33]. It may also be that elderly people, who are not very ill and not confined to their homes, decided not to consume the medical services they know and are insured with, as to not burden the medical staff who were stretched thin, as well as their fear of being exposed to those routinely working with sick people. Thus, the elderly avoided going to hospitals and being examined, especially those with chronic medical conditions, and preferred to deal with and treat themselves. This phenomenon is observed in times of crisis when people who are in distress avoid calling security services and try to fend for themselves [25]-[27].

The elderly’s calls to the center during the COVID-19 period decreased, and on the other hand, the average number of ambulances calls and a doctor’s home visits increased. In addition, the average number of service refusals increased compared to the previous year. In light of these findings, it can be assumed that the decrease in the average medical service calls may be due to the elderly’s attempts to manage on their own, and those who were unable to manage, having no other choice, seeking medical services such as an ambulance or a doctor’s home visit. Calling for a doctor’s home visit is an attempt to avoid arriving at an emergency room and being exposed to a greater number of people thus endangering themselves with unnecessary exposure to patients.

Older people require more medical services than young people [14]. The current study also found that the average service requirement among older adults over the age of 85 is higher. This is also the age group that refused more of the services. Moreover, the widowed subscribers consumed on average more services than single and divorced individuals, and significantly more than married ones. In this study, marital status was found to be a substantial and significant predictor for medical services demand, with the likelihood of unmarried people to require medical services being higher, as is supported by the literature [18], [20], [21].

When examining the literature to explore consumption of medical services by gender, it is shown that women more than men, tend to consume more medical services, adhere more to medical guidelines, obtain more of the required tests, and arrive more at the emergency room for medical treatments [10], [15]-[17]. Similarly, the study findings support the hypothesis and show that women in both time periods consumed significantly more medical services than men. The decrease that occurred in the demand for services during the COVID-19 lockdown, pertains to both sexes.

Ethnic gaps between Jews and Arabs may stem, among other things, both from cultural differences as well as from accessibility to health centers [14]. In the USA, higher rates of COVID-19 morbidity have been observed among ethnic minorities, whom have less access to medical services and health promotion abilities and have a lower socio-economic status relative to the general population. Minorities have less access to medical care and have a higher incidence of underlying morbidity leading to more serious disease [28]. This contrasts with our study data which shows that subscribers living in non-Jewish or mixed localities consumed on average more services than subscribers living in Jewish localities. Perhaps this is different in Israel where there is public health system to all and the demand for services is more accessible to vulnerable populations than in the USA. In addition, subscribers living in localities ranked in a lower than average or average socio-economic status according to the CBS [31] consumed more medical services compared to subscribers living in localities ranked in above-average socioeconomic status localities.

The logistic regression findings show that the most significant predictor of service demand during COVID-19 is service demand during the previous year, in a manner in which the chance of service demand during the first COVID-19 lockdown period is approximately 2.5times higher among those who required service in the previous year. This demonstrates the greater need that exists among at-risk populations, which require emergency medical services even during routine times.

The COVID-19 pandemic has significantly intensified the need for remote monitoring and treatment of patients, as a result of the patients’ fear of coming into contact with medical staff during the pandemic and has revolutionized the health care system in the field of telemedicine. Telemedicine kits are used among “Natali Healthcare Solutions in Israel” field workers to remotely monitor subscribers and document within their medical files, in a way that completely changes the medical approach preceding the pandemic outbreak [6]. The fear among seniors to receive essential medical care and their fear of contracting COVID-19 raises the vital need to use these systems for the service of the elderly in Israel.

The study’s findings indicate a decrease in medical service demand during COVID-19 which can be explained due to the elderly’s fear of being exposed to the disease through various service providers such as medical staff or other patients in different inpatient facilities, thus refraining from initially obtaining services. It is also possible that this concern dictated more refusals to receive a service offered by the call center operator. However, the increase in the demand for ambulance services and doctor’s home visits can be explained by the fact that when the subscribers did finally make the call for help, their situation was already more complex. Subscribers who did not go to the hospital on their own remained at home and were treated by various service providers as a substitute for hospital care. Another reason for the increase in the consumption of medical services is the reduction in the accessibility of community health services at the beginning of COVID-19. The Israeli health maintenance organizations tended to alienate patients from reaching the clinics due to the risk of infection. As alternatives, they suggested phone calls with the attending physician and receiving prescriptions remotely.

A very large sample of adults in Israel who consume medical services during routine times, is one of the main strengths of the present study. The data file of “Natali Healthcare Solutions in Israel”, a leading private company in Israel for providing medical care to the elderly, was analyzed here for the first time. Another strength of the study is its being a longitudinal study, based on the same subjects in both time periods, which contributes to the strength its findings. On the other hand, part of the study limitations is the lack of information pertaining to additional data such as the degree of religiosity, which could not be unequivocally examined since the data was based solely on place of residence, and not based on self-report. Another study limitation is that potential intervention variables such as disease severity, number of diseases, mental state, and nutritional balance, were not taken into account in this study, given the unavailability of this data.

The findings of the study show that there is a decrease in referrals to medical centers during times of emergency as well as during the first COVID-19 lockdown period, but an increase in the demand for medical services such as ambulance orders and doctor’s home visits as well as service refusals. There is no significant difference in the trends of the findings between the time periods according to different socio-demographic characteristics. The pandemic period raised concerns among the elderly about receiving medical care, possibly due to fear of contracting the disease, and was characterized by a higher incidence of medical service refusals during the pandemic.

Consumption of services among the elderly during COVID-19 decreased during the first lockdown compared to the previous year. The elderly’s fear of receiving services from an unfamiliar person / the fear of unnecessary exposure is known in times of crisis when people who are in distress avoid calling security services and try to fend for themselves [25]-[27]. Our study supportively showed that the elderly are afraid of going to the emergency room or receiving medical care and home care services during a pandemic. Therefore, policymakers in the health care system, in local authorities and in other government offices, that provide services to the elderly, must plan a tailored and better response to the needs of this population in times of a health crises.

In summary, this study is based on a large sample of elderly in Israel, findings indicate a decrease in referrals to medical care during the first COVID-19 lockdown period, yet an increase in ambulance orders, physician visits and service refusals. Moreover, the period of the pandemic was characterized by a higher incidence of medical service refusals compared to routine times. There were no significant differences between the periods according to different socio-demographic characteristics. The chance of requiring medical service at the time of the first COVID-19 lockdown was higher among those who required services in a previous year, among unmarried seniors, and among the older age group of 85 and over in comparison to the younger seniors. The implications of these findings reinforce the need for social response for the elderly in order to improve their quality of life, especially during a pandemic period in which adult life is at risk. Inter-institutional cooperation is required to produce good social programs for the preservation of the quality of lives of the elderly.

## Data Availability

All data produced in the present study are available upon reasonable request to the authors

## Acknowledgements

We would like to express our sincere gratitude to Mr. Nimrod Altman, “Natali Healthcare Solutions Israel” CEO for his kindness in providing the company database of subscribers.

## Notes

### Competing Interest Statement

The authors have declared no competing interest.

### Funding Statement

This study did not receive any funding

### Author Declarations

The study was approved by the Ariel University institutional ethics committee (Approval No. AU-HEA-AZ-20200624) and has also received the approval of "Natali Healthcare Solutions in Israel" management to conduct the research

